# A Dual-Strategy Roadmap to qMSP Reference Optimization: Boosting Efficiency and Pass Rates in Clinical Samples

**DOI:** 10.1101/2025.08.25.25334047

**Authors:** Shan Zhang, Yixuan Tang, Jiahui Jiang, Ling Chen

## Abstract

**Objective:** This study aimed to identify the optimal internal reference primer-probe set for multiplex qMSP, based primarily on amplification efficiency, to improve the detection pass rate for tumor DNA methylation analysis in low-DNA-concentration samples.

**Methods:** Our screening strategy comprised three steps. First, we used SYBR Green-based qPCR to preselect primer pairs with robust amplification. Second, we applied probe-based qPCR to further evaluate these candidates and identify superior primer-probe combinations. Finally, we compared the amplification performance of the finalists across diverse sample types, including cell lines, tissues, blood cells, and plasma, to determine the optimal set.

**Results:** Two optimal primer-probe sets were successfully identified: a performance-optimized set (bisACTB_103_F2+R2+P2) and a novel set (bisProAB1_F2+R2+P1). Both demonstrated superior amplification efficiency across various sample types. Crucially, the bisProAB1_F2+R2+P1 set significantly enhanced the amplification of low-concentration plasma cell-free DNA, thereby increasing the detection pass rate for minimal DNA input samples.

**Conclusion:** This study provides two highly efficient internal reference systems for qMSP. The novel bisProAB1_F2+R2+P1 set, in particular, offers a substantial improvement for detecting low-concentration clinical samples, enabling better utilization of precious biospecimens.

## Introduction

DNA methylation biomarkers are of significant value for the early screening and diagnosis of human cancers^1-3^。Quantitative methylation-specific PCR (qMSP) is a predominant technique for their detection, primarily utilizing two chemistries: the dye-based method (e.g., SYBR Green) and the probe-based method (e.g., TaqMan hydrolysis probes)^4^. This technique targets bisulfite-converted DNA (bisDNA), which is derived from human genomic DNA (gDNA) through sodium bisulfite treatment. Currently, most commercially available DNA methylation detection kits employ probe-based qPCR.

qMSP has been extensively applied to detect various malignancies, including colorectal cancer^5,6^, head and neck cancer^7^, lung cancer^8^, liver cancer^9,10^, breast cancer^11,12^, gynecological cancers^13,14^, and urothelial carcinoma^15,16^. The biospecimens analyzed encompass diverse clinical sample types such as stool^17^, saliva^7,18^, bronchoalveolar lavage fluid (BALF)^19^, tissue^20,21^, cervical exfoliated cells^14^, urine^15,16^, and blood (plasma/serum)^21-25^. In the aforementioned studies, the Cycle threshold (Ct) value range from the reference gene channel in qMSP assays is commonly used as a criterion to assess both DNA yield and sample adequacy. Similarly, commercial kits define sample acceptability based on a predetermined reasonable Ct range for the reference channel in probe-based qPCR. However, existing reference primer-probe sets frequently suffer from suboptimal amplification efficiency, resulting in elevated Ct values. This can lead to the misclassification of adequate samples as inadequate, causing wastage of valuable clinical specimens. Furthermore, the performance of reference primer-probe combinations reported in the literature and used in commercial kits is highly variable. Critically, there is a notable lack of systematic studies dedicated to optimizing and comparatively evaluating reference systems for qMSP.

This study was designed to overcome these limitations via a dual-strategy optimization framework. First, we refined primer-probe combinations targeting the *ACTB* reference gene, which have been frequently employed in studies analyzing colorectal cancer stool samples, esophageal cancer blood/plasma specimens, and liver/colorectal cancer blood samples. Through sequence optimization, we developed improved primer-probe sets that demonstrate enhanced amplification efficiency for bisulfite-converted DNA (bisACTB) while significantly reducing non-specific amplification of unconverted genomic *ACTB* (gACTB). Second, we systematically screened the human *ACTB* gene and its 2000 bp upstream promoter region to identify three CpG-free sequences as candidate targets. Based on the bisulfite-converted sense strand sequences of these regions, novel reference primer-probe sets were designed, several of which exhibited excellent amplification performance for bisACTB. These findings offer considerable value to both research and clinical communities by providing high-performance qMSP reference sets that enable significantly improved detection pass rates for clinical samples with low DNA concentrations.

## 1. Materials and Methods

### 1.1 Human Specimens

All samples (tissue and blood) were residual material obtained following the completion of routine clinical diagnostic procedures. Specifically, the paired cancer tissues and adjacent normal tissues consisted of archived frozen remnants from standard pathological examinations, while the preoperative peripheral blood samples, obtained from patients with benign gynecological conditions or malignancies, were residual aliquots collected in EDTA-K2 anticoagulant tubes for standard clinical testing; these samples were sourced specifically from the Department of Gynecology at Zhangjiajie People’s Hospital.

### 1.2 Cell lines

The HeLa, SKOV3, and IOSE80 cell lines were purchased from Procell Life Science & Technology Co., Ltd (Cat#CL-0101 and CL-0215; Wuhan; China) and Wuhan Sunncell Biotechnology Co., Ltd (Cat# SNL-626), respectively.

### 1.3 Primers and Probes Design

All primers were designed using Primer Premier 5 software with melting temperatures (Tm) of 58∼62℃. Probes were designed with Primer Express 3.0.1, featuring 5’-VIC fluorophore and 3’-MGB quencher modifications (Tm: 68∼73℃).

### 1.4 NCBI Primer-BLAST Analysis

Primer specificity was evaluated using NCBI Primer-BLAST (https://www.ncbi.nlm.nih.gov/tools/primer-blast/), where forward and reverse primers were entered in their respective fields. The analysis was performed using the "Genomes for selected eukaryotic organisms (primary assembly only)" database with Homo sapiens specified as the target organism, while maintaining default settings for the "Get Primers" function. The output results provided comprehensive information including genomic specificity profiles and amplicon size distributions.

### 1.5 DNA Extraction and Bisulfite Conversion

Genomic DNA from high-DNA-concentration samples (cell lines, blood cells, and tissues) was extracted using the TIANamp Genomic DNA Kit (DP304-03; TIANGEN, Beijing, China). Subsequently, 2 μg of genomic DNA (in ≤30 μL volume) was subjected to bisulfite conversion using a column-based methylation bisulfite conversion kit (Cat#122-002; GeneTech, Shanghai, China), followed by quantification of bisulfite-converted DNA on a NanoDrop™ One spectrophotometer (840-317400; Thermo Fisher Scientific). For low-DNA-concentration samples, plasma DNA was extracted with a magnetic bead-based plasma DNA extraction kit (N902-02; Vazyme Biotech, Nanjing, China), and 20 μL of the extracted DNA was converted using a magnetic bead-based methylation bisulfite conversion kit (Cat#EM103-01; Vazyme Biotech, Nanjing, China). All procedures were performed in accordance with the manufacturers’ protocols.

### 1.6 SYBR Green qPCR

The amplification performance of primer pairs was evaluated using three control configurations: (1) negative control with 1 ng/μL genomic DNA from HeLa cells (gHela), (2) positive control with 0.5 ng/μL bisulfite-converted DNA from HeLa cells (bisHela), and (3) no-template control with ddH□O. All qPCR reactions were performed in triplicate in a 20 μL mixture consisting of 10 μL of 2 × T5 Fast qPCR Mix (SYBR Green I; Cat# TSE202, Tsingke Biotechnology, Beijing, China), 0.4 μL each of forward and reverse primers (10 μM; synthesized by Sangon Biotech, Shanghai, China), and 2 μL of template DNA. Amplification was carried out on a SLAN-96S real-time PCR system (Hongshi Medical Technology, Shanghai, China) using the following thermal protocol: initial denaturation at 95°C for 1 min, followed by 45 cycles of 95°C for 15 s and 60°C for 15 s, with fluorescence acquisition during the 60°C step. A melting curve analysis was subsequently performed by gradually increasing the temperature from 60°C to 95°C at a rate of 0.03°C per second with continuous fluorescence monitoring.

### 1.7 Probe-Based qPCR

The amplification performance of primer-probe combinations were validated using the same negative and positive controls as previously described. Each 25 μL qPCR reaction contained 0.1 μL of Hot-start Taq DNA polymerase (antibody modified; Cat#HMD0206, Hzymes Biotechnology, Wuhan, China), 2.5 μL of 10× reaction buffer, 0.25 μL each of forward primer, reverse primer, and probe (10 μM; primers synthesized by Sangon Biotech Co., Ltd., Shanghai, China; probe synthesized by Bailige Biotechnology Co., Ltd., Shanghai, China), and 2 μL of template DNA. The reactions were run on a SLAN-96S real-time PCR system (Hongshi Medical Technology, Shanghai, China) under the following thermal protocol: 25°C for 10 min; 95°C for 2 min; 50 cycles of 95°C for 15 s and 60°C for 20 s, with fluorescence acquisition during the 60°C step. For sample testing, either bisulfite-converted DNA diluted to 0.5 ng/μL (for cells, tissues, and blood cells) or 2 μL of undiluted plasma-derived bisDNA was used as template, with all reactions performed in triplicate.

### 1.8 Data Visualization

All figures were created using GraphPad Prism 9.0.0 and Adobe Illustrator 2021 SP. Amplification curves from both SYBR Green assays (including melting curves) and probe-based qPCR assays were exported from real-time PCR instruments and plotted with uniform axis scales.

### 1.9 Statistical Analysis

Intergroup comparisons used two-tailed Student’s *t*-tests, with statistical significance set at P < 0.05.

## 2. Results

### 2.1 Dual-Strategy Development of a Universal qMSP Internal Reference via Three-Stage Screening

We employed two complementary strategies to optimize the internal reference system for qMSP (Figure 1). Strategy 1 focused on refining previously published primer-probe combinations to enhance their performance. Strategy 2 involved the design of entirely novel internal reference primers and probes; recognizing that the *ACTB* genomic regions used in prior literature all contained at least one CpG site, we targeted CpG-free regions within the ACTB gene and its promoter for new assay development. Both strategies adhered to a consistent three-stage screening pipeline. In the first stage, SYBR Green qPCR was used to identify optimal primer pairs based on two primary criteria: amplification efficiency, prioritized by exponential amplification curves, lower Ct values, and a single melting peak with Tm ≥ 80°C; and specificity, assessed by the absence or minimal presence of primer-dimer (melting peak < 80°C) or non-specific product (melting peak > 85°C) signals, with late-cycle amplification (Ct ≥ 40) being tolerated for negligible interference. The second stage utilized TaqMan probe-based qPCR to evaluate the top primer pairs combined with probes, selecting combinations that exhibited superior amplification efficiency (characterized by exponential curves and lower Ct values) and optimal probe performance (indicated by a strong fluorescence signal and low background). In the final stage (generalisable validation), the leading candidates were compared against each other across a diverse panel of sample types—including human cancer cell lines, normal cell lines, cancer tissues and matched adjacent normal tissues, blood cells and plasma from cancer patients, and counterparts from patients with benign diseases—to determine the most robust and universally applicable primer-probe combination.

**Figure 1.**
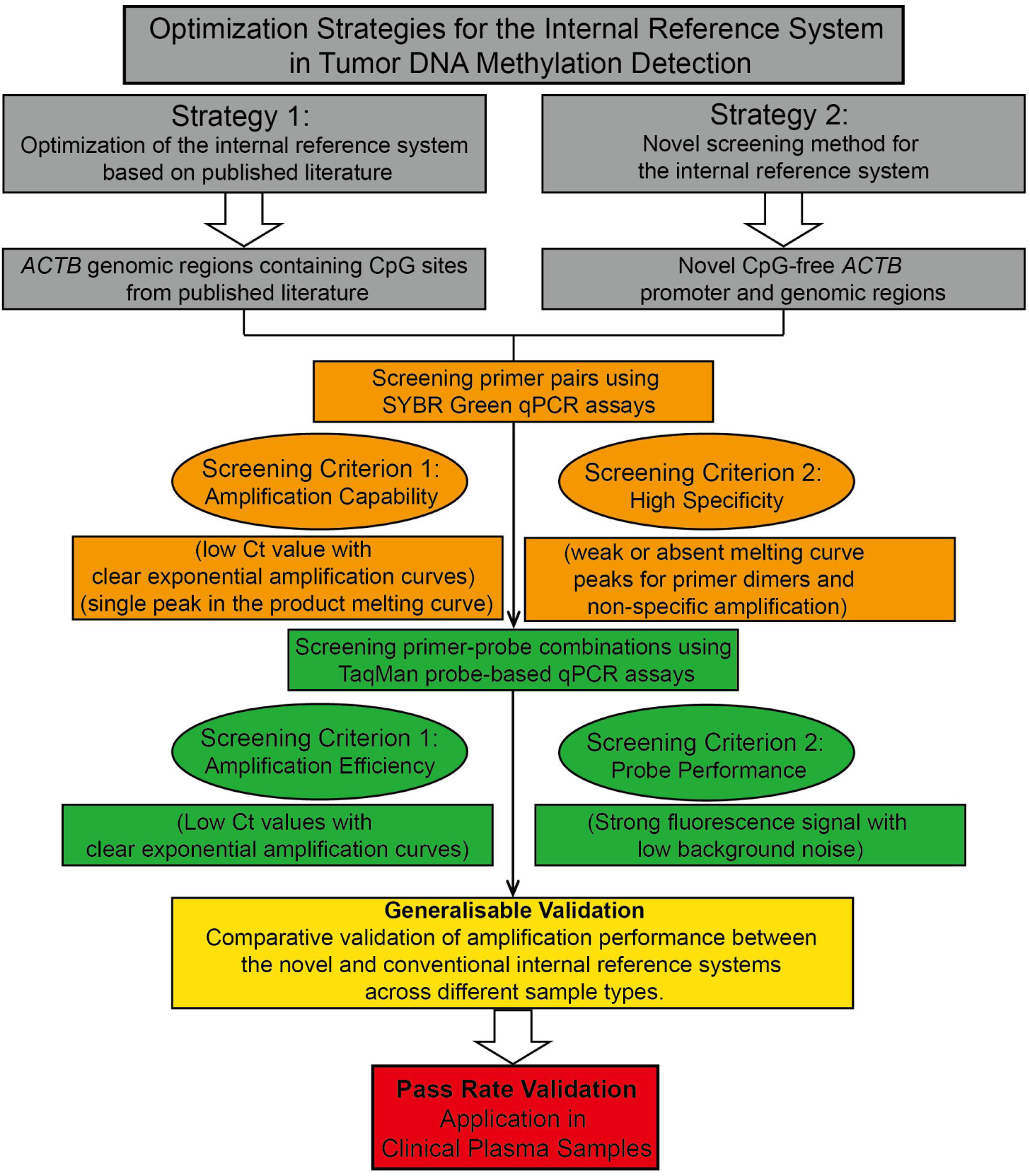
The Dual-Strategy Development of a Universal qMSP Internal Reference via Three-Stage Screening.

### 2.2 Systematic SYBR Green Optimization of Reported *ACTB* Primers Eliminates gDNA Interference and Enhances Bisulfite-Specific Amplification

Housekeeping genes, such as *ACTB*, are commonly used as internal reference controls in qMSP for tumor studies. Based on primer sequences targeting the region gACTB_103 (containing 2 CpG sites; see Supplementary Sequence Information) reported by Zhang et al.^26^, Wang et al. ^27^, and Bian et al.^28,29^, we designated this primer pair as bisACTB_103_F1 (F1: AAGGTGGTTGGGTGGTTGTTTTG) and bisACTB_103_R1 (R1: AATAACACCCCCACCCTGC), though sequence analysis revealed an A-to-G mutation at the second base from the 3’ end in R1 (Supplementary Sequence Information). To address potential limitations, we optimized the primers: Firstly, F1 was shortened by 2 nucleotides to bisACTB_103_F2 (F2: GTGGTTGGGTGGTTGTTTTG), reducing Tm from 64.8°C to 58.3°C. Secondly, R1 was extended by 4 nucleotides (avoiding the 3’ mutation) to bisACTB_103_R2 (R2: TTTTAATAACACCCCCACCCTAC), maintaining Tm near 59.5 ° C. Additionally, mutant bisACTB_103_F2_17C (F2_17C: GTGCTTGGGTGGTTGTTTTG) was designed to reduce dimerization with R2.

Incomplete bisulfite conversion (<100% efficiency) results in residual gDNA, which acts as a potent source of non-specific amplification in qMSP. Consequently, SYBR Green I-based qPCR was utilized to simultaneously assess three key parameters: (i) amplification efficiency of the bisACTB target, quantified using Ct values; (ii) product specificity evaluated through melting curve analysis for both bisDNA and gDNA templates; and (iii) primer-dimer artifact formation in no-template controls (NTCs). For this purpose, gDNA extracted from HeLa cells (gHeLa) and its bisulfite-converted counterpart (bisHeLa) served as template sources, while nuclease-free water functioned as the blank control (BC). Results demonstrated that F1+R1 amplified BC (Ct=33.55, melt peak 75-80 °C; Supplementary Figure 1A), indicating primer dimers. Although specific amplification was observed for both gHeLa (Ct=25.62; Tm=85-90°C) and bisHeLa (Ct=29.67; Tm=80-85°C; Supplementary Figure 1A,D; Table 1), the substantially lower Ct value for gHeLa indicated significant contamination risk from unconverted DNA. In contrast, optimized pair F2+R2 produced only primer-dimer peaks when amplifying gHeLa (Supplementary Figure 1B), while exhibiting markedly reduced Ct values for bisHeLa (25.82 vs. 29.67; p<0.01, Supplementary Figure 1D,E; Table 1), demonstrating elimination of gDNA amplification and enhanced bisACTB targeting efficiency. Furthermore, F2_17C+R2 displayed negligible dimer formation (undetectable in BC, Supplementary Figure 1C), absence of gDNA amplification products, and superior bisHeLa amplification efficiency (Ct=27.72; Supplementary Figure 1D,F; Table 1). Computational validation via NCBI Primer-BLAST identified a 103-bp genomic target for F1+R1, whereas no homologous targets were predicted for optimized primer pairs (Supplementary Sequence Information), corroborating experimental findings. Collectively, these results establish that both optimized primer pairs (F2+R2 and F2_17C+R2) effectively mitigate gDNA interference and improve bisACTB amplification, with F2_17C+R2 exhibiting minimal dimerization and thus representing an optimal internal reference for SYBR Green I-based qMSP in DNA methylation studies.

Subsequent optimization targeted the *ACTB* primer pair reported by Li et al. ^30^and Zhao et al. ^31^, which amplifies a bisACTB_89 fragment (89 bp; GC content 38-40%; containing 2 CpG sites, Supplementary Sequence Information). The original primers were designated as bisACTB_89_FP (FP: CGCAATAAATCTAAACAAACTCC), bisACTB_89_RP-1 (RP-1: GGGTTAGATGGGGGATATGT), and bisACTB_89_RP-2 (RP-2: AGGTTAGACGGGGGATATGT). To enhance thermodynamic properties, FP was extended by one nucleotide to generate bisACTB_89_R (R: CCGCAATAAATCTAAACAAACTCC), increasing its Tm from 56.4°C to 59.8°C, while RP-1/RP-2 were extended by five nucleotides to create bisACTB_89_F (F: CAGTTAGGTTAGATGGGGGATATGT), elevating Tm from 55.2°C/57.9°C to 60.2°C. Comparative qPCR analysis revealed that the original primer combinations (FP+RP-1 and FP+RP-2) exhibited inefficient bisHeLa amplification (Ct = 33.32 and 32.39, respectively; Supplementary Figure 2D,E; Table 3) despite showing no detectable amplification with gHeLa or blank controls (Supplementary Figure 2A,B,D,E). Conversely, the optimized F+R pair demonstrated significantly improved amplification efficiency for bisHeLa (Ct = 29.11; Supplementary Figure 2F; Table 3) without generating non-specific products (Supplementary Figure 2C,F). Although NCBI Primer-BLAST analysis indicated no predicted genomic amplicons for either primer set, the optimized F+R pair provides substantially enhanced bisACTB amplification efficacy, establishing its utility as a robust internal reference for qMSP assays.

Finally, we optimized the *ACTB* primer pair reported by Wang et al.^32^, targeting a bisACTB_78 fragment (78 bp; GC content 37-38%; containing 1 CpG site, Supplementary Sequence Information). The original primers, designated bisACTB_78_F2 (F2: GGGATATGTAGAAAGTGTAAAG) and bisACTB_78_R2 (R2: CACAATAAATCTAAACAAACTCC) (Supplementary Figure 3A,C), were modified as follows: F2 was extended by three nucleotides to generate bisACTB_78_F3 (F3: TGGGGGATATGTAGAAAGTGTAAAG; Tm increased from 48.7°C to 59.2°C) (Supplementary Figure 3B,E) and its mutant derivative bisACTB_78_F3_19C (F3_19C: GGGGGCTATGTAGAAAGTGTAAAG; Tm 59.8°C), while R2 was substituted with the previously optimized bisACTB_89_R (Tm increased from 51.6°C to 59.8°C) (Supplementary Figure 3C,F). qPCR evaluation demonstrated that the original F2+R2 combination exhibited inefficient bisHeLa amplification (Ct = 38.55; Supplementary Figure 3D; Table 5), whereas the optimized F3_19C+R pair achieved significantly enhanced amplification efficiency (Ct = 30.32; Supplementary Figure 3F; Table 5) without detectable amplification in gHeLa or blank controls (Supplementary Figure 3C,F). Critically, F3_19C+R produced a single melting curve peak for bisHeLa (Tm 75-80°C; Supplementary Figure 3C), consistent with expected product characteristics. Bioinformatic analysis indicated three low-significance genomic targets for F2+R2 via NCBI Primer-BLAST, while no comparable targets were predicted for optimized primers (data not shown). Consequently, F3_19C+R demonstrates high-specificity bisACTB amplification with undetectable gDNA interference and negligible dimer formation, establishing it as a high-fidelity internal reference for SYBR Green I-based qMSP in DNA methylation studies.

### 2.3 Systematic Probe-based Optimization of Reported *ACTB* Primer-probe Combinations Enhances Amplification Efficiency, Signal-to-Noise Ratio, and Specificity

Minor groove binder (MGB)-modified probes are widely employed in TaqMan probe-based qPCR for DNA methylation analysis due to their high specificity, shorter sequence requirements, and low fluorescent background^33-35^. Accordingly, all TaqMan qPCR assays in this study utilized MGB probes. However, since the probe sequence (GGAGTGGTTTTTGGGTTTG) used by Zhang et al.^26^, Wang et al.^27^, and Bian et al.^28,29^ initiates with a guanine (G) at its 5’-end—a suboptimal configuration for MGB probes—we redesigned a universal probe sequence designated bisACTB_103_P2 (abbreviated P2: AAACCTCCAAACAACC). This modification avoids a 5’-terminal G while positioning the probe closer to the 3’-end of the reverse primer than in published designs, thereby theoretically enhancing 5’ nuclease cleavage efficiency by Taq polymerase and improving signal-to-noise ratios. TaqMan qPCR results demonstrated that optimized primer-probe sets F2+R2+P2 and F2_17C+R2+P2 achieved significantly lower Ct values for bisHeLa amplification (28.33 and 28.54, respectively) compared to F1+R1+P2 (30.35) (Supplementary Figure 4A; Supplementary Table 2). Critically, no amplification was observed for gHeLa with either optimized set, whereas F1+R1+P2 showed partial amplification in replicates (Supplementary Figure 4B; Supplementary Table 2). All sets exhibited no amplification in blank controls (BC) (Supplementary Figure 4C; Supplementary Table 2). Collectively, these results confirm enhanced bisACTB targeting efficiency and absence of gDNA interference with optimized primer-probe combinations.

For the ACTB primer pairs from Li et al.^30^and Zhao et al.^31^, which shared the probe sequence TCCCAAAACCCCAACACAC, we designated a universal probe bisACTB_89_P (abbreviated P). TaqMan qPCR analysis revealed no significant difference in bisHeLa amplification efficiency between original (FP+RP-1+P and FP+RP-2+P) and optimized (F+R+P) primer-probe sets (Supplementary Figure 4D; Supplementary Table 4). Moreover, no amplification occurred with gHeLa or BC templates, indicating equivalent performance between literature-derived and optimized configurations in TaqMan assays.

Regarding the probe sequence (CCCAACACACTTAACCA) used by Wang et al.^32^with *ACTB* primers—designated bisACTB_78_P2 (P2)—its distal positioning relative to the 3’-end of bisACTB_78_R2 theoretically reduces Taq polymerase exonuclease activity, yielding suboptimal signal-to-noise ratios. To address this, we substituted P2 with bisACTB_89_P (P), which resides closer to the reverse primer’s 3’-end, and paired it with optimized primers F3+R and F3_19C+R. Notably, the original F2+R2+P2 set exhibited low plateau signal intensity, poor signal-to-noise ratio, and non-canonical amplification curves for bisHeLa (Supplementary Figure 4G). In contrast, optimized sets F3+R+P and F3_19C+R+P displayed significantly higher plateau signals and signal-to-noise ratios with canonical exponential curves (Supplementary Figure 4G), validating that proximity between the probe’s 5’-end and primer’s 3’-end enhances detection sensitivity. While Ct values showed no significant differences across sets (Supplementary Figure 4G; Supplementary Table 6), confirming comparable amplification kinetics, all sets demonstrated equivalent specificity with no amplification in gHeLa or BC samples (Supplementary Figure 4H,I).

Collectively, we established two optimized internal reference systems: (i) bisACTB_103_F2 + bisACTB_103_R2 + bisACTB_103_P2, which demonstrated significantly enhanced amplification efficiency compared to literature-reported configurations; (ii) bisACTB_78_F3 + bisACTB_89_R + bisACTB_89_P, which exhibited substantially improved amplification kinetics and signal-to-noise ratios. Both systems serve as robust internal references for TaqMan probe-based qPCR in tumor DNA methylation studies.

### 2.4 De Novo Design and Systematic Screening Establish Optimal Primer-Probe Sets for CpG-Free BisACTB Sequences

Since the gACTB fragments (gACTB_103, ACTB_89, and ACTB_78) referenced in prior studies contain ≥1 CpG site (Supplementary Sequence Information), bisulfite conversion may introduce T/C heterogeneity at these positions, resulting in sequence non-uniqueness (Supplementary Sequence Information). This variability could impair sample identification reliability when amplifying internal reference targets. To circumvent this limitation, we selected two CpG-free gACTB sequences and one *ACTB* promoter fragment^36^, designated gAB1, gAB2, and gProAB1 (Supplementary Sequence Information). Their bisulfite-converted counterparts (bisAB1, bisAB2, bisProAB1) exhibit sequence uniqueness, for which we designed dedicated primers and probes (Supplementary Table 14). NCBI Primer-BLAST analysis confirmed no predicted human genomic targets of comparable size for these novel primer pairs (data not shown).

SYBR Green qPCR revealed broad or multi-peak melting curves for bisAB1/bisAB2 primer pairs during bisHeLa amplification (data not shown); conversely, three bisProAB1 primer pairs— F+R^36^ (133 bp; 33% GC), F1+R1 (64 bp; 38% GC), and F2+R2 (78 bp; 35% GC)—generated single, sharp melting peaks for bisHeLa (Tm 75-80°C; Supplementary Figure 5A), consistent with predicted characteristics. Although Ct values showed no significant differences between F+R and F1+R1/F2+R2 (Supplementary Figure 5D; Supplementary Table 7), all sets exhibited primer dimers or non-specific amplification with gHeLa (Supplementary Figure 5B,E), and critically, only F+R lacked dimer peaks in blank controls (Supplementary Figure 5C,F). Thus, these reference sets are unsuitable for SYBR Green I-based tumor DNA methylation studies.

Given these limitations, TaqMan qPCR was employed to identify optimal configurations: For bisAB1, F4+R4+P4 demonstrated the lowest Ct (29.56) with canonical sigmoidal curves (Supplementary Figure 6A,D,G) and no gHeLa/BC amplification (Supplementary Figure 6B,C,E,F,H,I; Supplementary Table 8); for bisAB2, F4+R4+P4 showed suboptimal kinetics (non-sigmoidal curve, Ct=29.81; Supplementary Figure 6J) despite no gHeLa/BC amplification (Supplementary Figure 6K,L; Supplementary Table 8); whereas for bisProAB1, F2+R2+P1 achieved the minimal Ct (28.74; Supplementary Figure 7A; Supplementary Table 9) among tested combinations with undetectable background interference (Supplementary Figure 7B,C; Supplementary Table 9). Consequently, experimental validation established F4+R4+P4 (bisAB1), F4+R4+P4 (bisAB2), and F2+R2+P1 (bisProAB1) as optimal primer-probe sets.

In summary, through systematic screening of novel primers and probes targeting CpG-free bisACTB sequences derived from gACTB fragments, we established validated optimal primer-probe sets for two bisACTB segments and one bisProACTB segment.

### 2.5 Comprehensive Evaluation Across Sample Types Identifies ProAB1_F2+R2+P1 as the Optimal Internal Reference for Both High and Low DNA Input

To comprehensively evaluate the performance of optimized primer-probe configurations, we established the following comparative groups from Strategy 1: (1) original bisACTB_103_F1+R1+P2 (abbreviated as 103_F1+R1+P2) versus optimized bisACTB_103_F2+R2+P2 (abbreviated as 103_F2+R2+P2) (Figure 2A, E); (2) original bisACTB_89_FP+RP-1+P and bisACTB_89_FP+RP-2+P (abbreviated as 89_FP+RP-1+P and 89_FP+RP-2+P, respectively) versus optimized bisACTB_89_F+R+P (abbreviated as 89_F+R+P) (Figure 2B, F); (3) original bisACTB_78_F2+R2+P2 (abbreviated as 78_F2+R2+P2) versus optimized bisACTB_78_F3+bisACTB_89_R+bisACTB_89_P (abbreviated as 78_F3+89_R+89_P) (Figure 2C, G); (4) original bisProAB1_F+R+P (abbreviated as ProAB1_F+R+P) versus three optimized candidates: bisProAB1_F2+R2+P1 (abbreviated as ProAB1_F2+R2+P1), bisAB1_F4+R4+P4 (abbreviated as AB1_F4+R4+P4), and bisAB2_F4+R4+P4 (abbreviated as AB2_F4+R4+P4) (Figure 2D, H).

**Figure 2.**
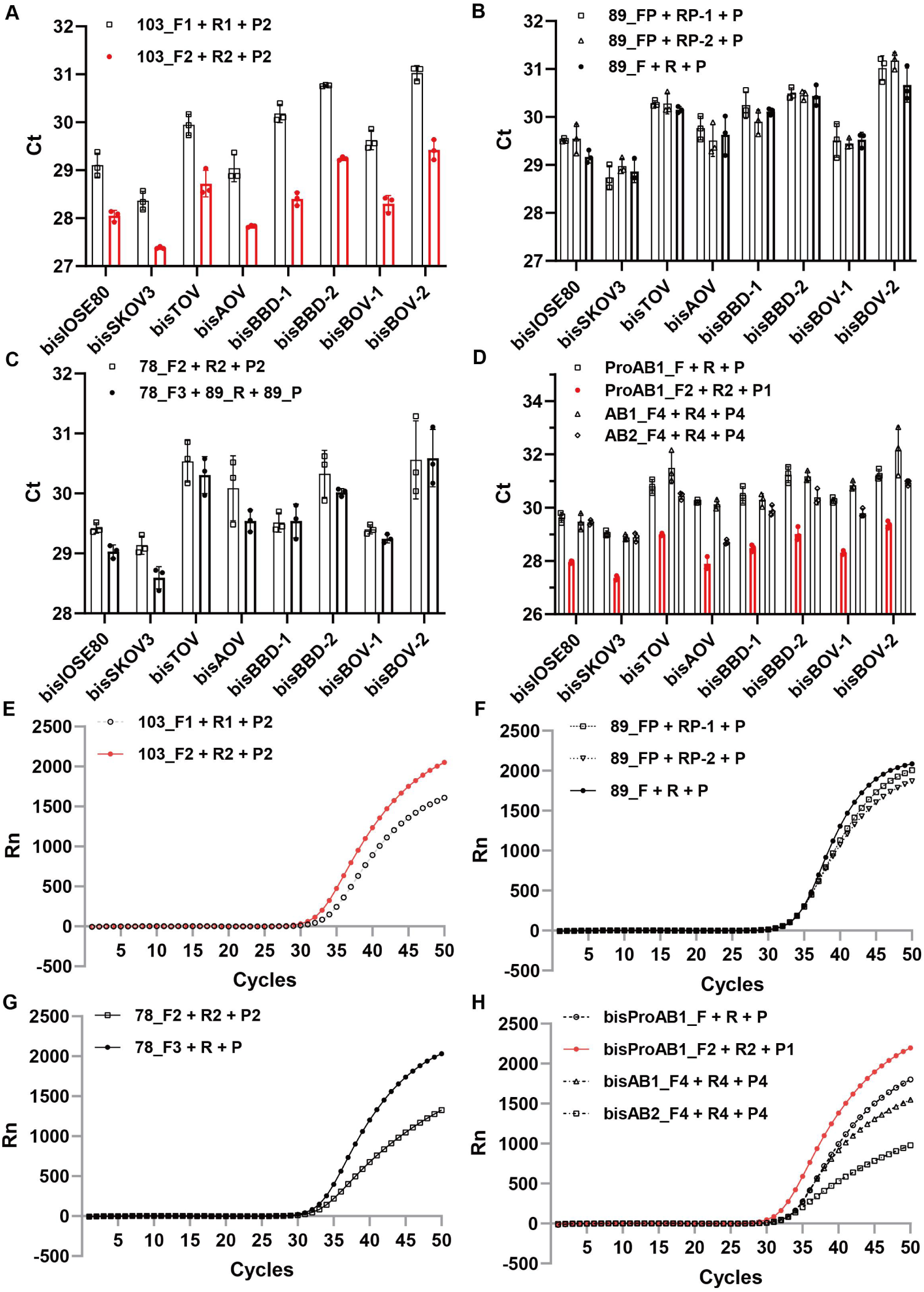
Comparison of Amplification Performance between Pre-optimization and Dual-Strategy-Optimized Primer-Probe Sets across Various High-DNA-Concentration Sample Types. (A–C) Amplification Ct values of primer-probe sets before and after optimization in Strategy 1 across various high-DNA-concentration sample types (bisIOSE80, bisSKOV3, bisTOV, bisAOV, bisBBD, bisBOV). Data points highlighted in red indicate one of the optimal primer-probe combinations (bisACTB_103_F2 + R2 + P2). (D) Amplification Ct values of the optimal primer-probe set from Strategy 2 compared to the control set across the same panel of high-DNA-concentration samples. The red data points denote one of the top-performing combinations (bisProAB1_F2 + R2 + P1). (E–G) Amplification curves of representative samples amplified using primer-probe sets before and after optimization in Strategy 1. Curves marked in red correspond to one of the optimal combinations (bisACTB_103_F2 + R2 + P2). (H) Amplification curves of representative samples using the optimal primer-probe set from Strategy 2 versus the control set. The red curve represents one of the best-performing combinations (bisProAB1_F2 + R2 + P1).

TaqMan probe-based qPCR was subsequently performed using bisDNA from high-DNA-concentration samples, including the IOSE80 and SKOV3 cell lines, ovarian carcinoma tissues (bisTOV), adjacent normal tissues (bisAOV), blood cell pellets from patients with benign disease (bisBBD), and those from ovarian cancer patients (bisBOV). The results demonstrated that the amplification Ct values of 103_F2+R2+P2 and ProAB1_F2+R2+P1 were significantly lower than those of their respective controls across all the aforementioned sample types (Figure 2A, D). Moreover, these two combinations exhibited superior probe performance, characterized by higher signal-to-noise ratios and improved amplification kinetics compared to their controls (Figure 2E, H). In contrast, no significant differences in Ct values were observed for 89_FP+RP-2+P and 78_F3+89_R+89_P relative to their corresponding controls across all high-DNA-concentration samples (Figure 2B, C). While the signal-to-noise ratio and amplification kinetics of 89_FP+RP-2+P showed no notable improvement over its control (Figure 2F), the 78_F3+89_R+89_P combination demonstrated significantly enhanced signal-to-noise ratio and amplification kinetics compared to its control (Figure 2G). Furthermore, when compared collectively against five original primer-probe sets, both 103_F2+R2+P2 (P = 5.4×10□¹²) and ProAB1_F2+R2+P1 (P = 2.5×10□¹²) exhibited significantly lower Ct values across all high-DNA-concentration samples (Figure 4A).

Plasma-derived cell-free DNA (cfDNA) represents a critical resource for discovering tumor DNA methylation biomarkers. However, its ultra-low concentration often precludes accurate quantification of bisDNA by conventional methods such as NanoDrop™. Therefore, establishing highly sensitive internal reference standards is essential for determining bisDNA content and improving sample qualification rates. To this end, we evaluated optimized primer-probe sets using bisDNA extracted from plasma samples of two representative patients with benign disease (bisPBD) and two with ovarian cancer (bisPOV). The results revealed that both 103_F2+R2+P2 and ProAB1_F2+R2+P1 exhibited significantly lower amplification Ct values compared to their respective controls in these low-DNA-concentration samples (Figure 3A, D). While the signal-to-noise ratio of 103_F2+R2+P2 showed no significant difference from its control (Figure 3E), ProAB1_F2+R2+P1 demonstrated superior signal-to-noise ratio and amplification kinetics over its control (Figure 3H). In contrast, 89_FP+RP-2+P and 78_F3+89_R+89_P showed no significant differences in Ct values relative to their controls (Figure 3B, C). Similarly, the signal-to-noise ratio and amplification kinetics of 89_FP+RP-2+P were not significantly improved compared to its control (Figure 3F), whereas 78_F3+89_R+89_P showed significantly enhanced signal-to-noise ratio and amplification kinetics (Figure 3G). Furthermore, when compared collectively against five original primer-probe sets, both 103_F2+R2+P2 (P = 5.4×10□□) and ProAB1_F2+R2+P1 (P = 2.5×10□¹¹) displayed significantly lower Ct values across all low-DNA-concentration samples. Notably, ProAB1_F2+R2+P1 demonstrated a higher degree of significance and greater Ct value stability (Figure 4B). In summary, ProAB1_F2+R2+P1 was identified as the optimal internal reference primer-probe combination.

**Figure 3.**
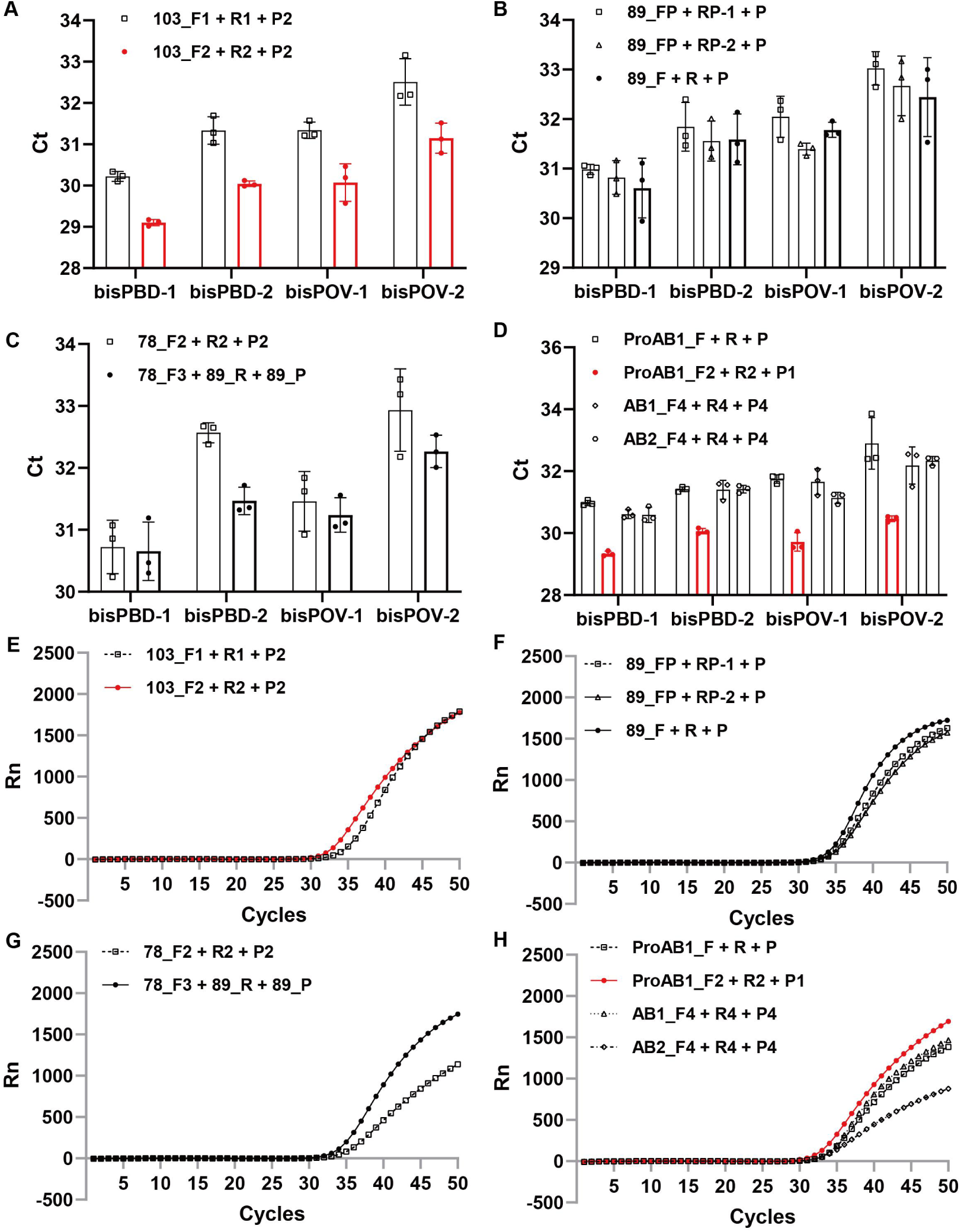
Comparison of Amplification Performance between Pre-optimization and Dual-Strategy-Optimized Primer-Probe Sets across Various low-DNA-Concentration Sample Types. (A–C) Amplification Ct values of primer-probe sets before and after optimization in Strategy 1 across various low-DNA-concentration sample types (bisPBD-1, -2; bisPOV-2, -2). Data points highlighted in red indicate one of the optimal primer-probe combinations (bisACTB_103_F2 + R2 + P2). (D) Amplification Ct values of the optimal primer-probe set from Strategy 2 compared to the control set across the same panel of low-DNA-concentration samples. The red data points denote one of the top-performing combinations (bisProAB1_F2 + R2 + P1). (E–G) Amplification curves of representative samples amplified using primer-probe sets before and after optimization in Strategy 1. Curves marked in red correspond to one of the optimal combinations (bisACTB_103_F2 + R2 + P2). (H) Amplification curves of representative samples using the optimal primer-probe set from Strategy 2 versus the control set. The red curve represents one of the best-performing combinations (bisProAB1_F2 + R2 + P1).

**Figure 4.**
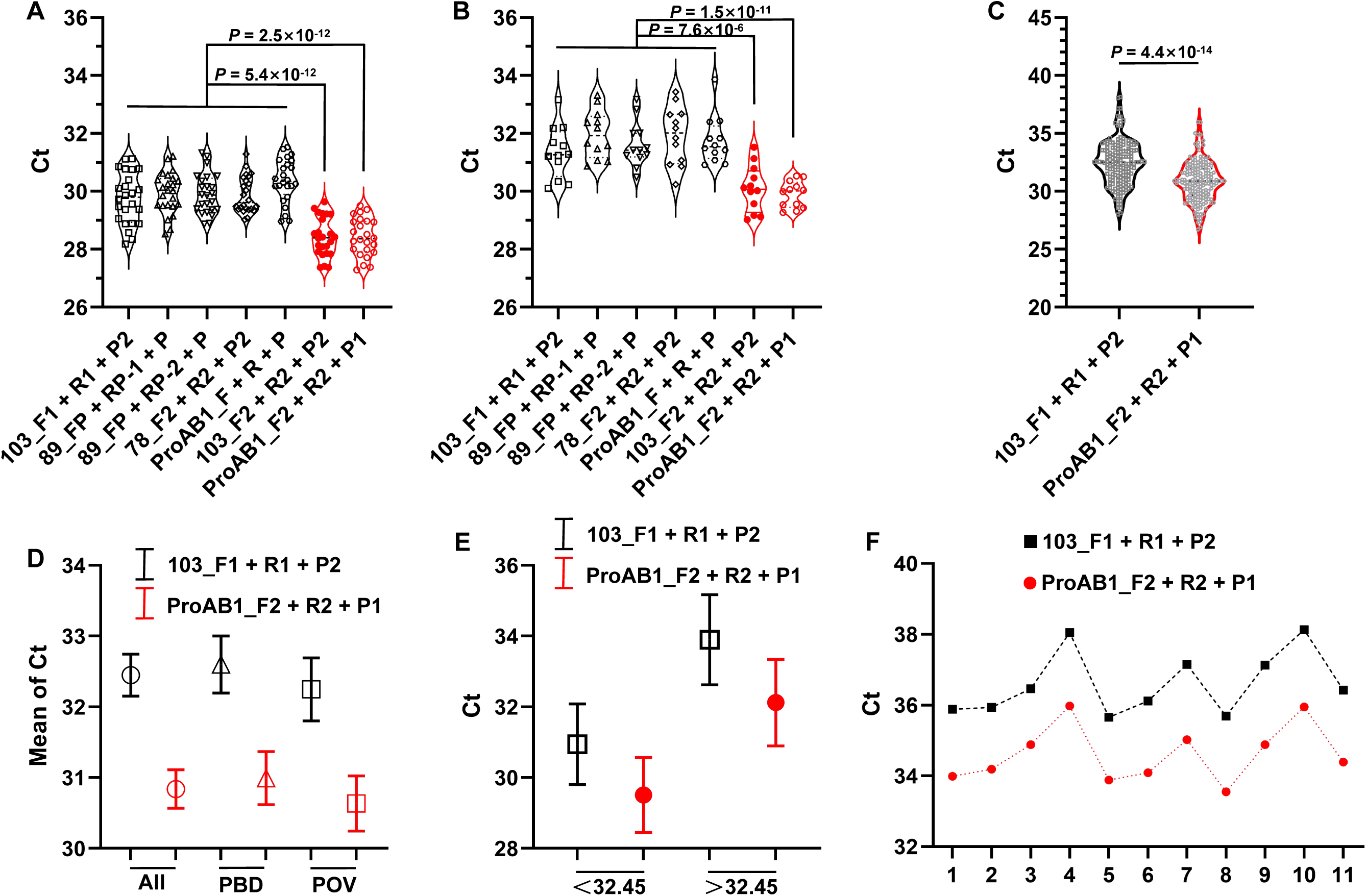
Identification of the optimal internal reference primer–probe combinations and its impact on detection pass rates in real-world plasma samples. (A) Amplification performance of the two top-performing primer–probe sets compared to five control sets in high-DNA-concentration samples. (B) Amplification performance of the two top-performing sets versus five control sets in low-DNA-concentration samples. (C) Comparison of amplification performance between the optimal primer–probe set (ProAB1_F2 + R2 + P1) and the control set (103_F1 + R1 + P2) across a large cohort of plasma samples. (D) Mean Ct values of the optimal set (ProAB1_F2 + R2 + P1) and the control set (103_F1 + R1 + P2) for different plasma samples. (E) Improvement in amplification efficiency conferred by the optimal set (ProAB1_F2 + R2 + P1) over the control set (103_F1 + R1 + P2), stratified by samples with Ct values above or below the mean. (F) Reduction in Ct values achieved by the optimal set (ProAB1_F2 + R2 + P1) compared to the control set (103_F1 + R1 + P2) in extremely low-DNA-concentration samples (Ct ≥ 35).

### 2.6 Novel Internal Reference Boosts Plasma Sample Pass Rate by ∼5% through Enhanced Detection Sensitivity

To investigate whether the optimal primer-probe combination (ProAB1_F2+R2+P1) could improve the detection pass rate for actual low-DNA-concentration plasma samples, we selected a previously reported primer-probe set (103_F1+R1+P2), which exhibited relatively better performance in earlier studies, as the control. Both the new and control sets were used to assess DNA quality in a large cohort of plasma samples, including 92 from patients with benign gynecological diseases (bisPBD) and 69 from ovarian cancer patients (bisPOV). The results showed that ProAB1_F2+R2+P1 yielded significantly lower Ct values than 103_F1+R1+P2 (Figure 4C). Across all 161 samples, the mean Ct value was 30.84 for ProAB1_F2+R2+P1 compared to 32.45 for the control set, corresponding to a reduction of 1.61 Ct units (equivalent to an approximately 3-fold lowering of the detection limit; Figure 4D). This improvement was consistent in both the bisPBD and bisPOV subgroups, with the same 1.61 Ct reduction observed in each (Figure 4D). When samples were stratified based on the control’s mean Ct value (32.45), ProAB1_F2+R2+P1 reduced Ct values by an average of 1.43 (□2.7-fold lower detection limit) in the 79 samples with Ct < 32.45, and by 1.78 (□3.4-fold lower detection limit) in the 82 samples with Ct > 32.45 (Figure 4E). These results indicate that the improved detection performance of the new internal reference set is more pronounced in samples with lower DNA concentrations (Ct > 32.45). In qMSP assays, samples with internal reference Ct values > 35 are typically considered unqualified. Statistical analysis revealed that 11 samples exceeded this threshold with the control set, resulting in a pass rate of 93.17% (150/161). In contrast, only 3 samples exceeded Ct > 35 with ProAB1_F2+R2+P1, achieving a pass rate of 98.14% (158/161)—an improvement of nearly 5% (Figure 4F). Moreover, in these extremely low-concentration samples (Ct > 35), ProAB1_F2+R2+P1 further reduced the mean Ct value by 1.98, corresponding to an approximately 4-fold lowering of the detection limit.

## 3 Discussion

### 3.1 Beyond Synthetic Templates: The Critical Role of Biological Templates in Faithful qMSP Assay Optimization

This study utilized gDNA and bisDNA as reference templates — rather than synthetic gACTB/bisACTB plasmid constructs—for two critical reasons: First, synthetic templates only permit amplification efficiency assessment without evaluating specificity interference, whereas biological templates enable simultaneous evaluation of target amplification capacity and non-specificity exclusion. Second, different primer pairs amplify distinct internal reference fragments, employing artificially synthesized internal controls would introduce multiple variables, precluding direct performance comparisons between primer pairs. To implement this approach, we initially utilized gDNA and bisDNA from cervical carcinoma HeLa cells as unified templates for screening primer/probe sets via SYBR Green qPCR and probe-based qPCR. Optimized configurations were subsequently validated across diverse biospecimens — including IOSE80/SKOV3 cell lines, gender-stratified tumor/adjacent tissues, blood cells, and plasma — ultimately identifying the highest-performance primer-probe combinations.

In practical applications of tumor DNA methylation qPCR kits, authentic biospecimens— including bisDNA from cell lines, tumor/adjacent tissues, blood cells, and plasma—are used as positive/negative controls and test samples. Our study comprehensively incorporated these clinically relevant sample types. Through a dual-strategy optimization approach aimed at enhancing bisDNA amplification sensitivity and gDNA specificity (Figure 1), we successfully improved the detection qualification rate for low-DNA-concentration clinical specimens, particularly plasma, in probe-based qPCR assays. This optimization minimizes the waste of precious samples while potentially increasing the detection positivity rate of the kit. Furthermore, using a large cohort of real-world clinical plasma samples, we validated the detection performance of our optimized system, demonstrating its ability to improve the detection pass rate for samples with extremely low DNA concentrations—thus achieving the primary objective of this study. Looking forward, researchers, developers, and clinical laboratory professionals working in tumor DNA methylation detection can adopt the internal reference system established in this work to enhance detection pass rates for precious low-DNA-concentration samples and significantly reduce unnecessary waste.

### 3.2 A Primer Designer’s Imperative: Incorporating gDNA Interference Mitigation from the Outset

Following bisulfite conversion, complementary strands of human gDNA lose sequence reciprocity. Consequently, primers designed against either converted strand must prioritize mitigation of gDNA interference. Systematic NCBI Primer-BLAST analysis of literature-derived, optimized, and novel primer pairs revealed that only the bisACTB_103 primers from Zhang et al. ^26^, Wang et al.^27^, and Bian et al.^28,29^ exhibited perfect genomic matches to a 103-bp endogenous target. Dye-based qPCR validation confirmed significant gDNA amplification for this set, characterized by distinct melting peaks (85∼90℃; Supplementary Supplementary Figure 5A,D) and significantly lower Ct values versus bisDNA (Supplementary Table 1). Probe-based qPCR further demonstrated non-specific amplification with gDNA templates (Supplementary Figure 5B). Critically, given ubiquitous gDNA contamination in qMSP laboratories, such primer pairs risk generating false-positive signals in negative controls—thereby compromising bisulfite conversion efficiency validation. In contrast, novel primer pairs experimentally demonstrated undetectable gDNA amplification, confirming effective mitigation of genomic interference.

### 3.3 Proactive Selection of CpG-Free Regions: Ensuring Conversion-Independent Amplification

The gACTB fragments referenced in prior studies^26^ contain 1-2 CpG sites (Supplementary Sequence Information). While authors assumed these CpG sites remain unmethylated (designing bisACTB primers against T templates), emerging evidence indicates *ACTB* methylation occurs in pathologies like stroke^37^ and coronary heart disease^38^. In such cases, methylated cytosines resist conversion, creating sequence heterogeneity at CpG sites. Primers targeting unconverted T templates may fail to amplify these variants, compromising internal reference functionality and sample qualification rates. To address this, we selected two CpG-free gACTB fragments and one CpG-free *ACTB* promoter fragment (bisAB1/bisAB2/bisProAB1) as reference regions, ensuring conversion consistency. Future studies should screen additional CpG-free genomic regions to develop higher-performance reference systems.

### 3.4 MGB Probes Overcome Primer-Dimer Interference to Restore BisACTB qPCR Efficiency

In SYBR Green assays, control primers for bisACTB_89 (FP+RP-1/FP+RP-2) and bisACTB_78 (F2+R2) showed significantly higher Ct values versus optimized sets (Supplementary Supplementary Figure 6D-F; Supplementary Tables 3,5). Strikingly, probe-based qPCR revealed equivalent efficiency between control and optimized combinations when paired with MGB probes (Supplementary Figure 5D,G; Supplementary Tables 4,6). This suggests primer-dimers suppressed target amplification in dye-based assays, while MGB probes likely counteracted such interference — potentially through enhanced specificity — to restore amplification efficiency.

### 3.5 A ΔCt-Based Quality Framework to Accelerate the Translation of DNA Methylation Biomarkers

Through multi-parameter optimization — including Tm harmonization (58-62□), dimer-disrupting mutations, NCBI Primer-BLAST verification, and probe positioning adjustments—we developed reference sets achieving ΔCt = -1.5∼-2.0 versus controls (Figure 4). Since each ΔCt = -1 reflects 2-fold sensitivity gain, these high-efficiency references not only improve sample qualification rates but also establish performance benchmarks for biomarker assays. We propose a ΔCt-based quality metric: Biomarker primers performing within ±0.5 ΔCt of the reference are "qualified"; >+0.5 ΔCt indicates "substandard"; <-0.5 ΔCt denotes "excellent". Implementing such standardized evaluation frameworks will accelerate translational development of DNA methylation biomarkers across research, diagnostic, and clinical domains.

## 4. Summary

This study establishes a systematically optimized internal reference system for quantitative methylation-specific PCR (qMSP) to address the critical challenge of high sample failure rates in DNA methylation analysis, particularly for low-input clinical specimens such as plasma cell-free DNA. We implemented a dual-strategy optimization framework: first, refining previously reported primer-probe sets for the *ACTB* gene through sequence modification to enhance thermodynamic properties and specificity; second, designing novel primer-probe combinations targeting newly identified CpG-free regions within the *ACTB* gene and its promoter, thereby ensuring consistent amplification irrespective of methylation status. A rigorous three-stage screening pipeline was employed, progressing from SYBR Green-based primer preselection to TaqMan probe-based evaluation, culminating in validation across a diverse panel of biospecimens, including cancer cell lines, tissues, blood cells, and plasma from patients with benign diseases and ovarian cancer. This comprehensive approach identified two optimal primer-probe sets: an optimized version of a literature-derived set (bisACTB_103_F2+R2+P2) and a novel set (bisProAB1_F2+R2+P1). Both sets demonstrated superior amplification efficiency, minimal primer-dimer formation, and effectively eliminated non-specific amplification from unconverted genomic DNA, a common source of false positives. Crucially, in a large cohort of 161 clinical plasma samples, the novel bisProAB1_F2+R2+P1 set significantly outperformed conventional references, reducing the mean Ct value by 1.61 cycles (equivalent to a ∼3-fold increase in sensitivity) and elevating the sample detection pass rate from 93.2% to 98.1%—a nearly 5% absolute improvement. The enhancement was most pronounced in challenging, ultra-low-DNA-concentration samples. By providing a robust, high-efficiency reference system and a standardized ΔCt-based quality metric for future biomarker assays, this work directly facilitates more reliable detection and reduces precious sample waste, thereby accelerating the translational application of DNA methylation biomarkers in cancer diagnostics.

## Supporting information

All supplementary data

## Waiver of Clinical Trial Registration Statement

This observational study utilized residual biological samples and, as defined by the International Committee of Medical Journal Editors (ICMJE), did not qualify as a clinical trial; thus, prospective registration was not required. The research employed a retrospective, non-interventional design. All tissue and blood samples were residual specimens obtained solely after the completion of routine clinical diagnostic procedures. Specifically, tissue samples comprised archived frozen remnants from standard pathological examinations, while blood samples were residual aliquots collected in EDTA-K2 anticoagulant tubes during standard clinical testing. No additional interventions were performed on participants, nor were any samples collected specifically for this study. Importantly, investigators had no role in the original clinical sampling decisions. All analyses were conducted using coded samples and associated de-identified clinical data. Written informed consent was obtained from all individuals whose residual samples were used in this study. This secondary use of residual samples complies with relevant ethical guidelines, including the Declaration of Helsinki, and received formal approval from the Clinical Research Management Committee of Zhangjiajie People’s Hospital (Approval No: [LL-2025-0017]).

## Data availability

The data underlying this article are available within the article [and/or its supplementary materials] and will be shared upon reasonable request to the corresponding author.

