## Supplementary material for "A Dual-Strategy Roadmap to qMSP Reference Optimization: Boosting Efficiency and Pass Rates in Clinical Samples": All supplementary data: Supplementary Figures.pptx

### Slide 1
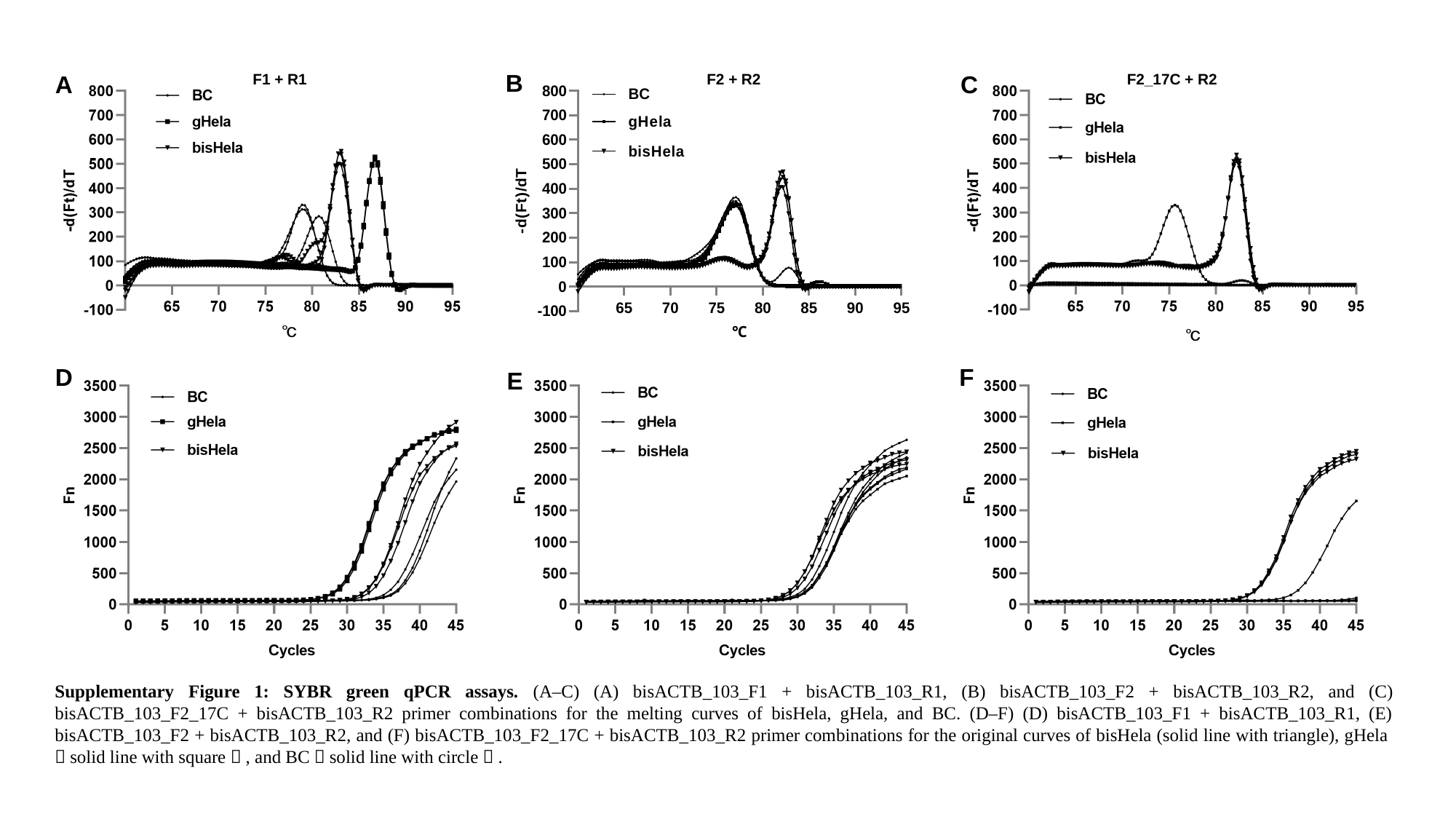

B
A
F1 + R1
F2 + R2
C
F2_17C + R2
D
F
E
Supplementary Figure 1: SYBR green qPCR assays. (A–C) (A) bisACTB_103_F1 + bisACTB_103_R1, (B) bisACTB_103_F2 + bisACTB_103_R2, and (C) bisACTB_103_F2_17C + bisACTB_103_R2 primer combinations for the melting curves of bisHela, gHela, and BC. (D–F) (D) bisACTB_103_F1 + bisACTB_103_R1, (E) bisACTB_103_F2 + bisACTB_103_R2, and (F) bisACTB_103_F2_17C + bisACTB_103_R2 primer combinations for the original curves of bisHela (solid line with triangle), gHela（solid line with square）, and BC（solid line with circle）.

### Slide 2
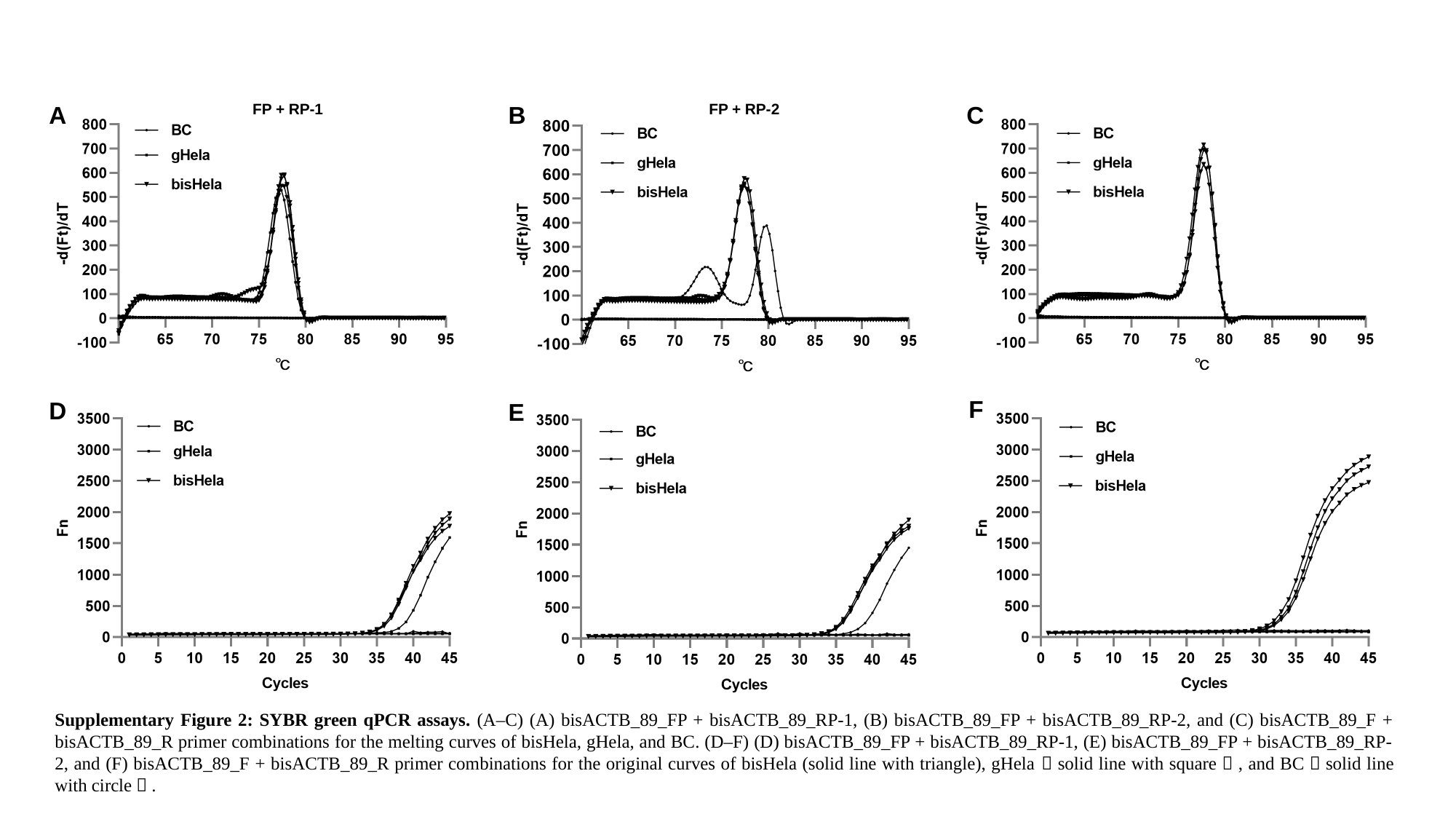

A
FP + RP-1
B
FP + RP-2
C
F
D
E
F + R
Supplementary Figure 2: SYBR green qPCR assays. (A–C) (A) bisACTB_89_FP + bisACTB_89_RP-1, (B) bisACTB_89_FP + bisACTB_89_RP-2, and (C) bisACTB_89_F + bisACTB_89_R primer combinations for the melting curves of bisHela, gHela, and BC. (D–F) (D) bisACTB_89_FP + bisACTB_89_RP-1, (E) bisACTB_89_FP + bisACTB_89_RP-2, and (F) bisACTB_89_F + bisACTB_89_R primer combinations for the original curves of bisHela (solid line with triangle), gHela（solid line with square）, and BC（solid line with circle）.

### Slide 3
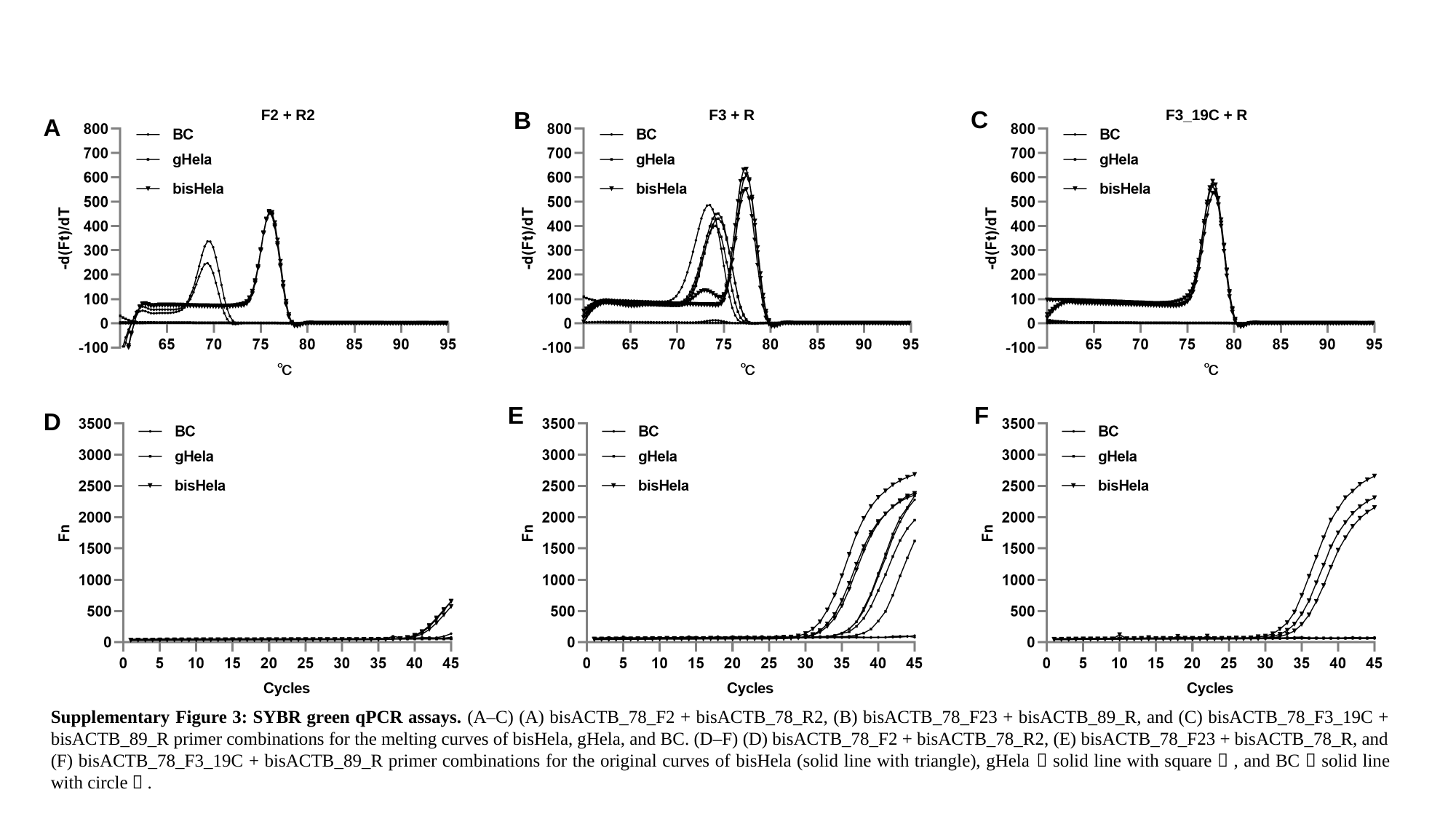

C
F2 + R2
B
F3 + R
F3_19C + R
A
E
F
D
Supplementary Figure 3: SYBR green qPCR assays. (A–C) (A) bisACTB_78_F2 + bisACTB_78_R2, (B) bisACTB_78_F23 + bisACTB_89_R, and (C) bisACTB_78_F3_19C + bisACTB_89_R primer combinations for the melting curves of bisHela, gHela, and BC. (D–F) (D) bisACTB_78_F2 + bisACTB_78_R2, (E) bisACTB_78_F23 + bisACTB_78_R, and (F) bisACTB_78_F3_19C + bisACTB_89_R primer combinations for the original curves of bisHela (solid line with triangle), gHela（solid line with square）, and BC（solid line with circle）.

### Slide 4
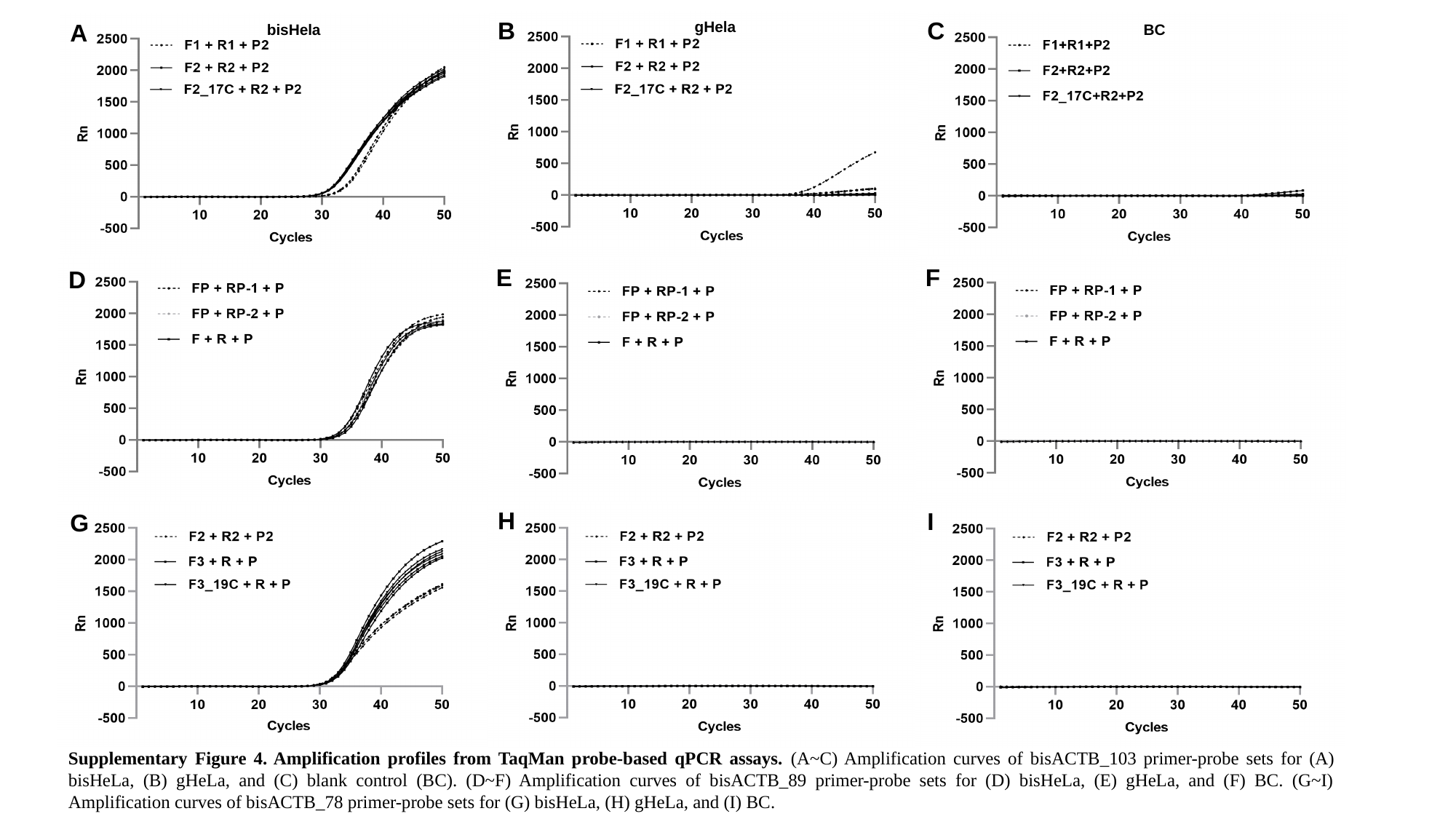

B
C
gHela
A
bisHela
BC
E
F
D
H
I
G
Supplementary Figure 4. Amplification profiles from TaqMan probe-based qPCR assays. (A~C) Amplification curves of bisACTB_103 primer-probe sets for (A) bisHeLa, (B) gHeLa, and (C) blank control (BC). (D~F) Amplification curves of bisACTB_89 primer-probe sets for (D) bisHeLa, (E) gHeLa, and (F) BC. (G~I) Amplification curves of bisACTB_78 primer-probe sets for (G) bisHeLa, (H) gHeLa, and (I) BC.

### Slide 5
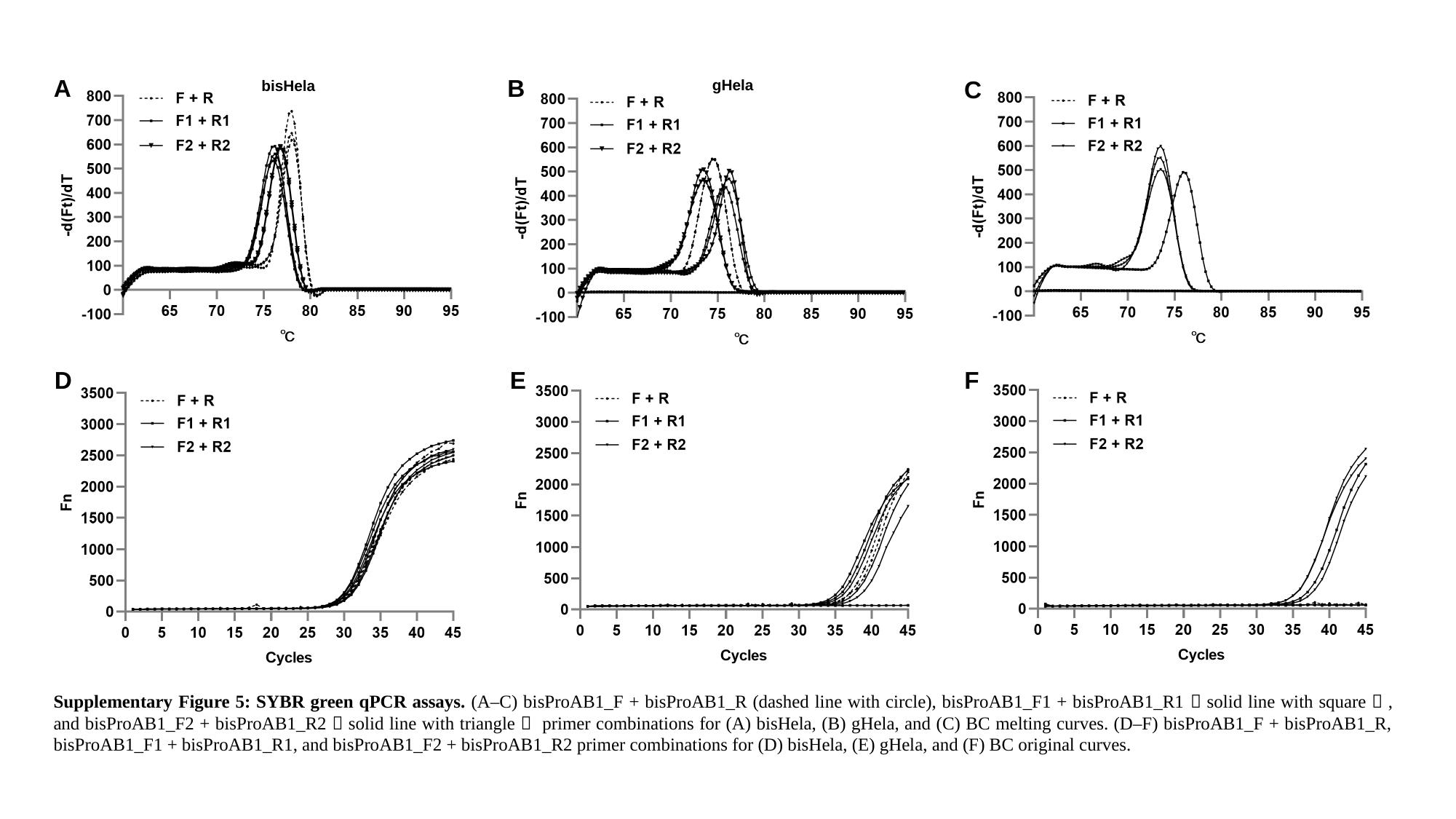

B
A
C
gHela
bisHela
E
D
F
BC
Supplementary Figure 5: SYBR green qPCR assays. (A–C) bisProAB1_F + bisProAB1_R (dashed line with circle), bisProAB1_F1 + bisProAB1_R1（solid line with square）, and bisProAB1_F2 + bisProAB1_R2（solid line with triangle） primer combinations for (A) bisHela, (B) gHela, and (C) BC melting curves. (D–F) bisProAB1_F + bisProAB1_R, bisProAB1_F1 + bisProAB1_R1, and bisProAB1_F2 + bisProAB1_R2 primer combinations for (D) bisHela, (E) gHela, and (F) BC original curves.

### Slide 6
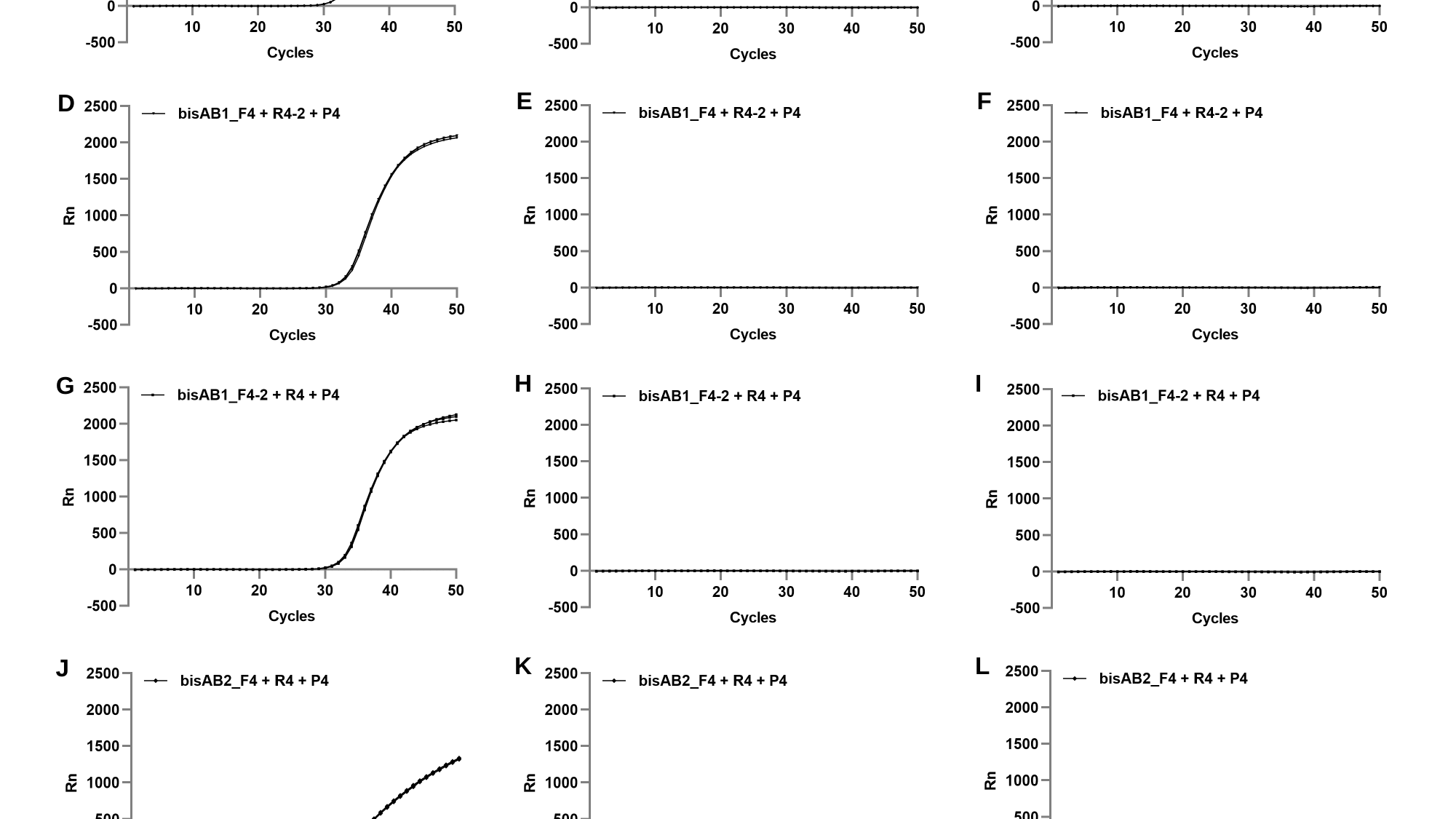

gHela
bisHela
BC
B
C
A
E
F
D
H
I
G
K
L
J
Supplementary Figure 6. Amplification profiles from TaqMan probe-based qPCR assays. (A-C) Amplification curves generated by the bisAB1_F4/R4/P4 primer-probe set for (A) bisHeLa (B) gHeLa, and (C) blank control (BC). (D-F) Amplification curves produced by the bisAB1_F4/R4-2/P4 combination for (D) bisHeLa, (E) gHeLa, and (F) BC. (G-I) Amplification profiles obtained with the bisAB1_F4-2/R4/P4 set for (G) bisHeLa, (H) gHeLa, and (I) BC. (J-L) Amplification curves using the bisAB2_F4/R4/P4 system for (J) bisHeLa, (K) gHeLa, and (L) BC.

### Slide 7
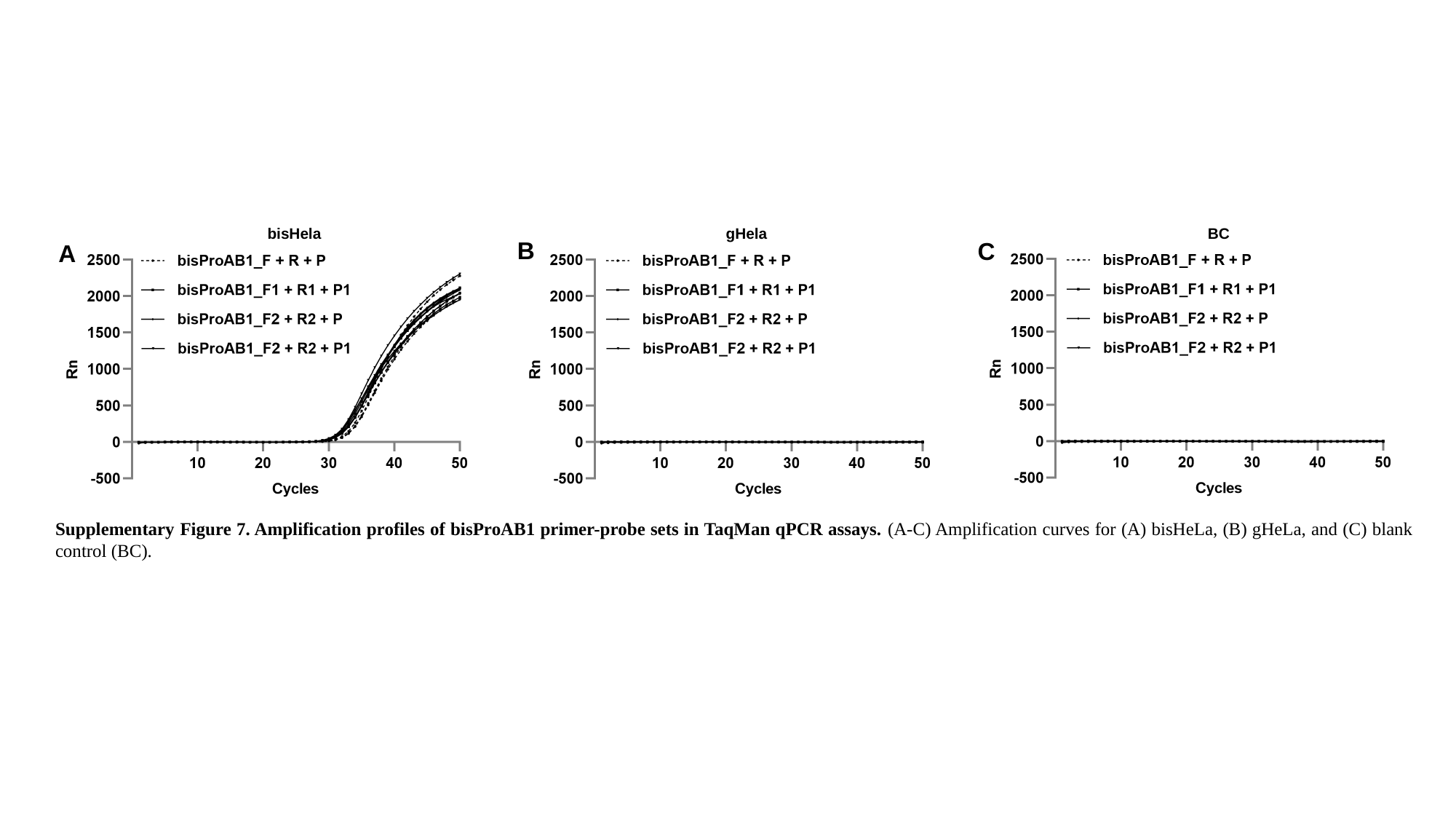

bisHela
gHela
BC
B
C
A
Supplementary Figure 7. Amplification profiles of bisProAB1 primer-probe sets in TaqMan qPCR assays. (A-C) Amplification curves for (A) bisHeLa, (B) gHeLa, and (C) blank control (BC).
