## Supplementary material for "A Dual-Strategy Roadmap to qMSP Reference Optimization: Boosting Efficiency and Pass Rates in Clinical Samples": All supplementary data: Supplementary sequence information.docx

**gACTB_103:** AAGGTGGCTGGGTGGTTGTTTTGCGGGAGGGCCAAGGAGTGGTCCCTGGGTCTGCGCTGTAAGAGTTGGTTGCCTGGAGGCCTTGCAGGGTGGGGGTGTCACC

**bisACTB_103:**

AAGGTGGTTGGGTGGTTGTTTTGCGGGAGGGTTAAGGAGTGGTTTTTGGGTTTGCGTTGTAAGAGTTGGTTGTTTGGAGGTTTTGTAGGGTGGGGGTGTTATT

**NCBI Primer-blast results of bisACTB_103_F1 and bisACTB_103_R1:**

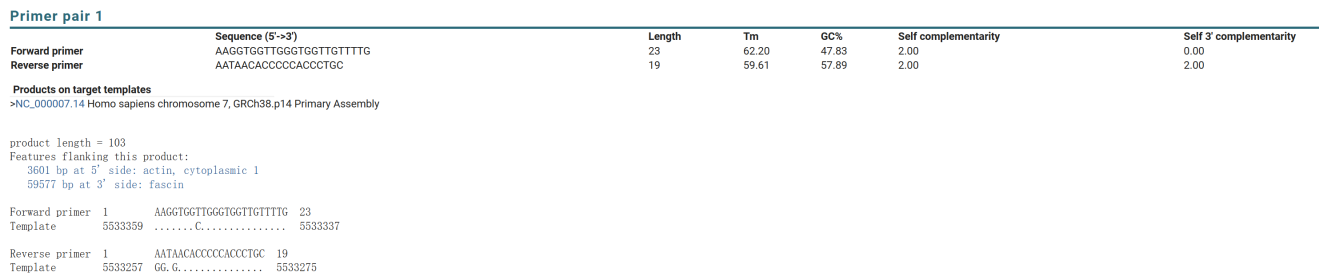

**NCBI Primer-blast results of bisACTB_103_F2 and bisACTB_103_R2:**

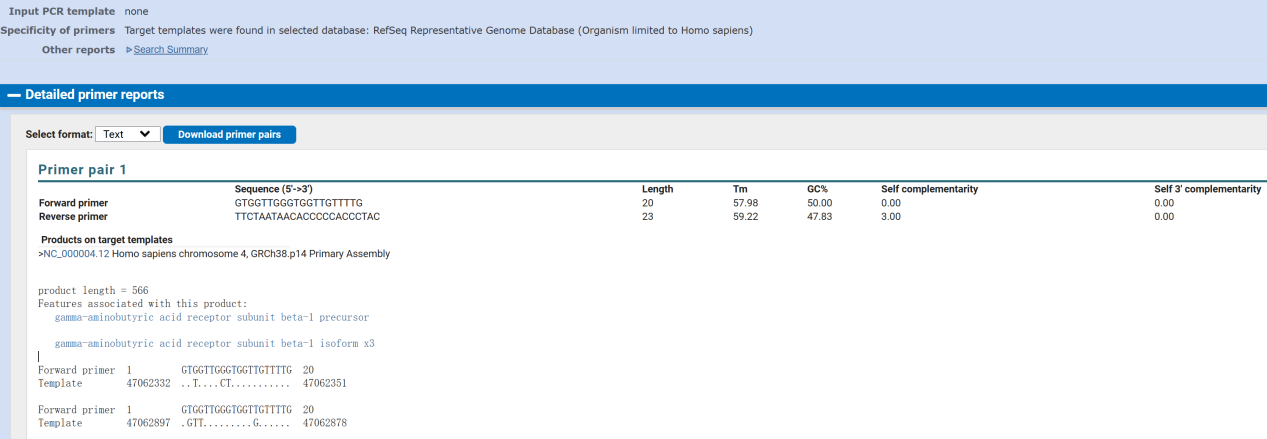

**gACTB_89:**

AGGCCAGACGGGGGACATGCAGAAAGTGCAAAGAACACGGCTAAGTGTGCTGGGGTCTTGGGATGGGGAGTCTGTTCAGACCTACTGTG

**bisACTB_89:**

AGGTTAGACGGGGGATATGTAGAAAGTGTAAAGAATACGGTTAAGTGTGTTGGGGTTTTGGGATGGGGAGTTTGTTTAGATTTATTGTG

**gACTB_78:**

GGGACATGCAGAAAGTGCAAAGAACACGGCTAAGTGTGCTGGGGTCTTGGGATGGGGAGTCTGTTCAGACCTACTGTG

**bisACTB_78:**

GGGATATGTAGAAAGTGTAAAGAATACGGTTAAGTGTGTTGGGGTTTTGGGATGGGGAGTTTGTTTAGATTTATTGTG

**gAB1:**

GGGTGTGGTGGTGCATGCCTGTGGTCCCAGCCACTTGGGAAACTGAGGTGGGAGGATTGCTCAAGCCCAGGAGGTCAAGGCTGCTGTGAGCTATGATTGTGACACTGCACTCCAGCCTGGGCAACAAAGGGAGACCCCTTAACTAACTAAATGAATGAATGAATGAATGAATGAAGGCAACCTGAGCTGCATTCCTTGGCCTGCCAACCTGCCCAGCCCCATCCCTCAGCCCTCCCTGAGTCTGAGGGCCCTGCAGGTCCCACACAGGGCCAGGCTCCATCTTGTTTCTGCAAATTTGCACCTTC

**bisAB1:**

GGGTGTGGTGGTGTATGTTTGTGGTTTTAGTTATTTGGGAAATTGAGGTGGGAGGATTGTTTAAGTTTAGGAGGTTAAGGTTGTTGTGAGTTATGATTGTGATATTGTATTTTAGTTTGGGTAATAAAGGGAGATTTTTTAATTAATTAAATGAATGAATGAATGAATGAATGAAGGTAATTTGAGTTGTATTTTTTGGTTTGTTAATTTGTTTAGTTTTATTTTTTAGTTTTTTTTGAGTTTGAGGGTTTTGTAGGTTTTATATAGGGTTAGGTTTTATTTTGTTTTTGTAAATTTGTATTTTT

**gAB2:**

GACATGCTTCCAGAGTTAACTGGCACTGTTTTGAGAAATGAGAAAAGCACCCTGAAGAAAGCAAGCACAATTCCTAAGACAGATCTTAAATGAGAGAGAGAGCAAGAGAACAACAAGTTTAATGGCATCAGGAGGAAAAGCAACAGGTTTCCTTCAGAATGAGGTATTAGGGGGTGCTGGGTGTAGTGGCTCATGCCTGTAATCCCAGCACTTTGGGAGGCTGAGGCAGGCAGATCA

**bisAB2:**

GATATGTTTTTAGAGTTAATTGGTATTGTTTTGAGAAATGAGAAAAGTATTTTGAAGAAAGTAAGTATAATTTTTAAGATAGATTTTAAATGAGAGAGAGAGTAAGAGAATAATAAGTTTAATGGTATTAGGAGGAAAAGTAATAGGTTTTTTTTAGAATGAGGTATTAGGGGGTGTTGGGTGTAGTGGTTTATGTTTGTAATTTTAGTATTTTGGGAGGTTGAGGTAGGTAGATTA

**gProAB1:**

TGGGGTGGTGATGGAGGAGGCTCAGCAAGTCTTCTGGACTGTGAACCTGTGTCTGCCACTGTGTGCTGGGTGGTGGTCATCTTTCCCACCAGGCTGTGGCCTCTGCAACCTTCAAGGGAGGAGCAGGTCCCATTGGCTGAGCACAGCCTTGTAC

**bisProAB1:**

TGGGGTGGTGATGGAGGAGGTTTAGTAAGTTTTTTGGATTGTGAATTTGTGTTTGTTATTGTGTGTTGGGTGGTGGTTATTTTTTTTATTAGGTTGTGGTTTTTGTAATTTTTAAGGGAGGAGTAGGTTTTATTGGTTGAGTATAGTTTTGTAT
